# The Impact of In Utero Tobacco Exposure on Cardiovascular Disease Risk and All-cause Mortality in Adulthood: a UK Biobank Study

**DOI:** 10.1101/2024.08.19.24312279

**Authors:** Yanxu Zheng, Xinyu Xiong, Jing Bao, Jingyu Liu, Jin Wang, Zixi Chen, Fang Zou, Yang Guo, Qingyao Wang, Yixuan Qiu, Zhaowei Zhu

## Abstract

**Aim:** The negative impacts of in utero tobacco exposure (IUTE) on cardiovascular disease (CVD) have been insufficiently described. This study aims to assess the association between IUTE and the risks of CVD incidence and all-cause mortality, discuss the inter-group difference based on genetic susceptibility and smoking behaviors after birth, and explore the potential mediating factors.

**Methods:** Utilizing a total of 375,024 participants from the UK Biobank, the outcomes include myocardial infarction, stroke, chronic ischemic heart disease, nonrheumatic aortic valve disorders, cardiomyopathy, heart failure, atherosclerosis, aortic aneurysm and dissection, and all-cause mortality.

**Results:** During a median follow-up period of 14.6 years, 50,434 cases of CVD were recorded. IUTE was significantly associated with increased CVD incidence (HR 1.10, 95% CI 1.08-1.12) and all-cause mortality (HR 1.11, 95% CI 1.09-1.14). Interaction effects between IUTE, smoking behaviors after birth, and genetic risk scores for CVD were observed significant (P for interaction < 0.005). The results of the cross-sectional study revealed a significant positive association between IUTE and smoking behaviors after birth (OR 1.08, 95% CI 1.06-1.09). Mediation analysis indicated that smoking behaviors (Proportion = 12.40%, P < 0.001) and HDL-c levels (Proportion = 14.20%, P < 0.001) partially mediated the IUTE-CVD relationship.

**Conclusions:** This study demonstrated that individuals with IUTE have a higher risk of developing CVD, and smoking behaviors after birth have multifaceted influence on this correlation. These findings underscore the importance of mothers avoiding smoking during pregnancy to mitigate adverse effects on their offspring.

## Introduction

In 2019, cardiovascular disease (CVD) was the primary cause of death and disability globally, accounting for approximately 18.6 million deaths and 393 million disability-adjusted life years [1, 2]. Given these staggering figures, there is a critical need for ongoing research to better understand the etiology, develop more effective treatments, and implement preventive strategies to reduce the global burden of CVD. Among all the heart-related health results studied so far, an increasing body of evidence suggests that apart from traditional CVD risk factors such as alcohol consumption, smoking, diabetes, and hypertension [3–6], there are intergenerational correlations between utero health during pregnancy and the risk factors of CVD in offspring [7–10].

According to a systematic review published in The Lancet Global Health, a significant portion of mothers—approximately 8% in the European region—smoke during pregnancy [11]. Research has consistently shown that smoking during pregnancy can negatively impact the health of an unborn child. A slew of studies has illustrated that in utero tobacco exposure (IUTE) might increase the likelihood of offspring developing asthma [12], accelerate biological aging in offspring during adulthood [13], contribute to the prevalence of childhood obesity [14], correlate with reduced brain volumes and suboptimal brain development in preadolescents [15], potentially increase the risk of mental health issues [16], and elevate the overall risk of psychiatric disorders in children [17]. Given the clear link between tobacco exposure and various adverse outcomes for children, it becomes evident that this issue extends far beyond individual choice. It represents a grave public health concern, bringing with long-term detrimental effects that reverberate through the lives of offspring [18, 19].

The existing body of research on the associations between intrauterine environmental factors (IUTE) and the full spectrum of CVD remains limited. Although previous studies have explored the relationship between IUTE and congenital heart defects in children, they face significant limitations. Notably, results from a Mendelian randomization study suggest that maternal smoking during pregnancy does not have a significant association with these outcomes [20]. Moreover, a scant number of follow-up studies have narrowly concentrated on certain types of CVD without a wider contextual analysis [21, 22]. Additionally, the apparent positive correlation highlighted in a parental negative control study seems to be predominantly influenced by a single case type [23], and collectively, these reports focus on congenital anomalies without contemplating the enduring cardiovascular repercussions in later life. Notably, a cohort study targeting various acquired CVDs in youth terminated at 18 years, an age not typically marked by high CVD prevalence [24]. This underscores the urgent need for more comprehensive research that includes older populations, who are more heavily affected by CVD, to clarify the potential long-term effects of tobacco exposure during pregnancy on the cardiovascular health of offspring. Accordingly, our study aims to meticulously assess the lasting impact of intrauterine tobacco exposure on the occurrence of various CVDs in the progeny during their middle to later years.

We utilized the cohort data of over 300,000 adults from the UK Biobank study to investigate the association between IUTE and CVD risks, discussed the potential intermediary factors, and explored the influence of genetic susceptibility.

## Methods

### Study participants

The UK Biobank is a large biomedical database and research resource containing de-identified genetic, lifestyle, and health information as well as biological samples from half a million UK participants (https://www.ukbiobank.ac.uk). It is a highly extensive and detailed prospective study that recruited over 500,000 participants aged between 40 and 69 years from 2006 to 2010. During this period, 22 assessment centers across the UK assessed the participants, covering a wide range of environments to provide socioeconomic, racial heterogeneity, and urban-rural mix. This ensured a broad distribution of all exposures, allowing for reliable identification of universal associations between baseline characteristics and health outcomes [25]. Ethical approval for the UK Biobank study was granted by the North West Multi-center Research Ethics Committee.

This study utilized resources from the UK Biobank (application number: 84709), with analysis restricted to a subset of participants with complete baseline data. Due to the close association of high-density lipoprotein cholesterol (HDL-c), low-density lipoprotein cholesterol (LDL-c), and triglycerides with the study outcomes and their considerable data missingness in the UK Biobank, we employed a Decision Tree model for multiple imputations. Using data from the remaining variables, we constructed decision trees to predict and impute missing values for these three variables. Detailed exclusion criteria are outlined in Figure 1. In summary, after imputation and exclusion, we included 351,392 participants without pre-baseline CVD to investigate the association between IUTE and CVD incidence, among whom 341,055 patients with complete standard PRS data were used to investigate the influence of genetic susceptibility. Additionally, 375,024 participants with complete death records were included in the study examining the association between IUTE and all-cause mortality. The cross-sectional analysis of IUTE and adverse behaviors utilized 375,024 participants with complete information.

**Figure 1.**
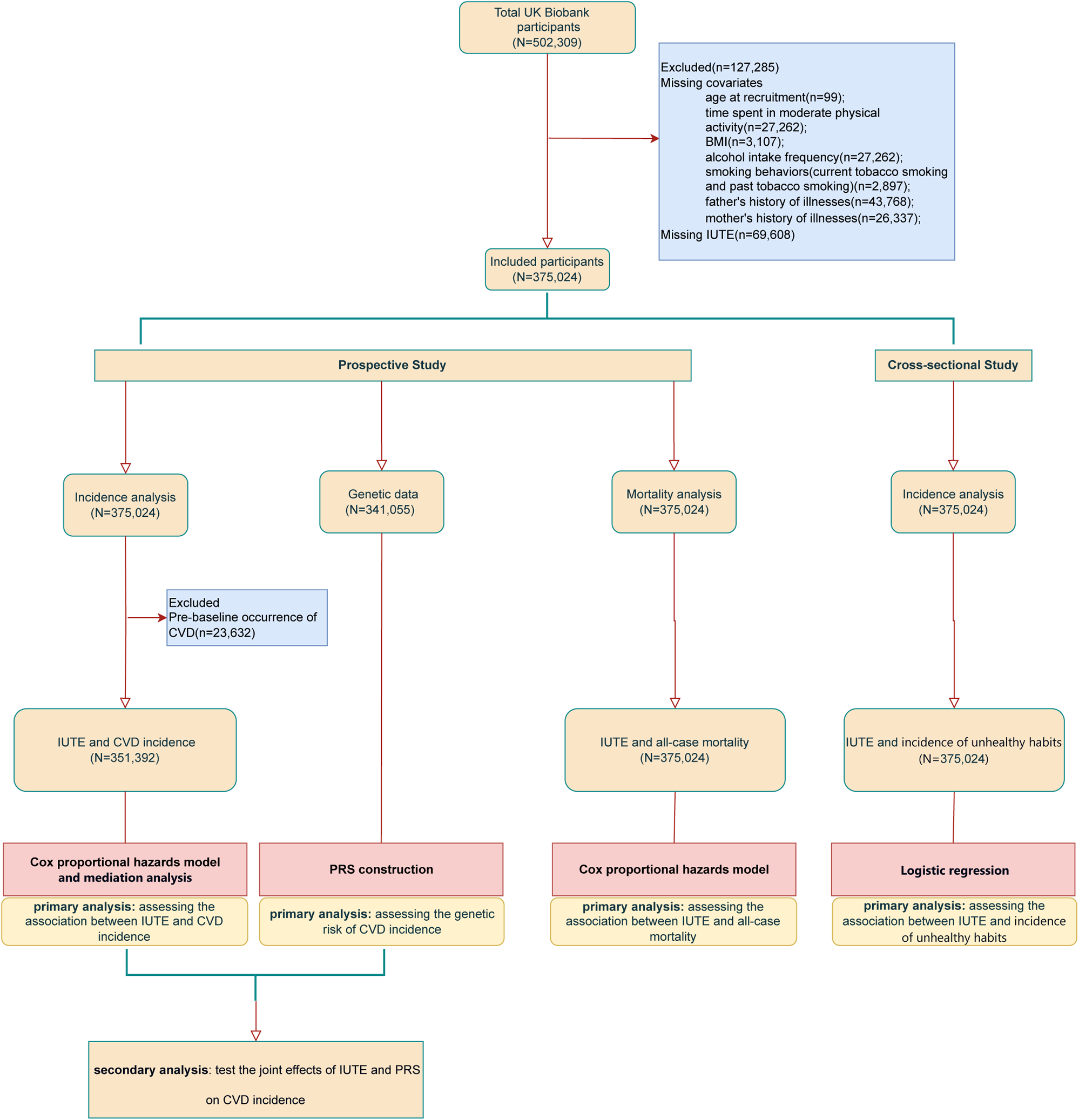
Flowchart of participants enrollment.

### Assessment of IUTE

IUTE was assessed based on the self-reported smoking status of the mother during pregnancy, obtained through touchscreen questionnaires at the UK Biobank assessment centers. Participants were categorized into two groups, “exposed to IUTE” and “unexposed to IUTE,” based on their responses.

### Outcome definition

The primary outcome events of this study are the incidence rates of eight clinically prevalent CVDs, classified using The International Classification of Diseases, Tenth Revision (ICD-10) coding (see Supplementary Table 1). Among them, records of the first occurrence of chronic ischemic heart disease, nonrheumatic aortic valve disorders, cardiomyopathy, heart failure, atherosclerosis, aortic aneurysm, and dissection are derived from primary care data, hospital inpatient data, death register records, and self-reported medical conditions at baseline or during follow-up visits to the UK Biobank assessment centers. Data on myocardial infarction and stroke are obtained by combining information collected from the UK Biobank baseline assessment, hospital admissions, diagnoses, procedures, and death registration records. “CVD” is defined as having any of the aforementioned eight diseases. Participants are followed up from enrollment until the first occurrence of the interested disease diagnosis, death, loss to follow-up, or the end of the follow-up date (September 30, 2023), whichever comes first. The secondary outcome event of the study is all-cause mortality. Death data on UK Biobank participants is received from NHS England for participants in England & Wales and from the NHS Central Register (NHSCR), part of the National Records of Scotland, for participants in Scotland.

### Assessment of covariates

Baseline population characteristics of patients were obtained from local NHS Primary Care Trust registries, including gender and age at enrollment. Time spent in moderate activity, ethnic background, alcohol intake frequency, smoking behaviors, father’s history of illnesses, and mother’s history of illnesses were derived from touchscreen questionnaires completed at the assessment centers during enrollment. HDL-c, LDL-c, and triglycerides were measured in blood samples collected at recruitment. Body mass index (BMI) was calculated as weight (measured using a BC-418MA body composition analyzer) divided by height (collected using a Seca 240cm stadiometer) squared (kg/m^2^). Smoking behaviors includes both current smokers and former smokers who have quit smoking. Drinking behaviors is categorized as never, occasionally (one to three times a month and special occasions only), and frequently (daily or almost daily, three or four times a week, and once or twice a week). The measure of time spent in moderate physical activities was documented by recording the number of days per week in which participants engaged in such activities for a minimum of 10 consecutive minutes each time. Ethnic background included the white race (British, Irish and any other white background) and others. The aforementioned covariates were included after being screened according to the following criteria: (1) the effect value of X on Y changed by more than 10% after adding the confounding factor; (2) previous studies have confirmed a significant association between this factor and the outcome.

### Acquisition of Polygenic Risk Score

To evaluate genetic susceptibility, PRS for CVD was extracted from the “Standard PRS” category provided by the UK Biobank (Data-Field 26223). This score, published by Thompson et al. in 2022 [26], was generated by meta-analyzing multiple external GWAS sources to produce a standard set of PRS for 28 diseases and 8 quantitative traits for all UKB participants (Category ID 301).

### Statistical analyses

Baseline characteristics of the study population were presented according to IUTE (yes or no). T-tests and chi-square tests were used to compare the characteristics between groups for continuous and categorical variables, respectively. We plotted Kaplan-Meier curves to illustrate the overall incidence of CVD over time and used the log-rank test to describe the differences between the IUTE group and the non-exposure group. Cox proportional hazard regression models with three adjustment levels were employed to calculate the hazard ratios (HR) and 95% confidence interval (95% CI) for the relationship between IUTE and the incidence of CVD as well as all-cause mortality. Model 1 was unadjusted for covariates; Model 2 adjusted for age at enrollment, gender, ethnic background, BMI, and time spent in moderate activity; Model 3 (the fully adjusted model) further adjusted for drinking behaviors, smoking behaviors, father’s history of illnesses, mother’s history of illnesses, HDL-c, LDL-c, and triglycerides.

Fully-adjusted (excluding smoking behaviors after birth) Cox proportional hazards model was applied in subgroups stratified by smoking behaviors after birth to determine the potential impact on the association between IUTE and CVD.

To evaluate the role of genetic susceptibility in the correlation between IUTE and the risk of CVD incidence, we grouped participants based on the quartiles of the standard PRS for CVD provided by UKB. We then conducted analyses within each group using fully adjusted Cox proportional hazards models.

The association between IUTE and adverse behaviors after birth (including smoking and drinking) was examined using cross-sectional analyses with logistic regression models. This analysis aimed to explore whether IUTE increases the likelihood of being exposed to tobacco and alcohol after birth, thereby potentially affecting their risk of developing CVD. Here, drinking behaviors was treated as a binary variable, classified as “Yes” (occasional drinking and weekly drinking) or “No” (never drinking). The three variable-adjusted models were as follows: Model 1 did not adjust for any covariates; Model 2 was adjusted for BMI, ethnic background and time spent in moderate activity; Model 3 (the fully adjusted model) further adjusted for drinking behaviors (only for the analysis of smoking), smoking behaviors (only for the analysis of drinking), HDL-c, and LDL-c.

In the mediation analysis, we investigated whether the association between IUTE and various CVD risks was mediated by smoking behaviors after birth and HDL-c. All pathways in the mediation analysis were evaluated by constructing Ordinary Least Squares Linear regression models. We utilized the Bootstrapping method by randomly sampling with replacement from the study sample 500 times to estimate the parameters and then averaged the results across each sampling to obtain the final estimation [27].

We conducted all statistical analyses using Visual Studio Code software (version 1.88.1 Universal). Basic data manipulation and analysis were performed using the pandas package. Survival analysis, including Kaplan-Meier and Cox proportional hazards models, was conducted using the lifelines package. Decision tree regression models and iterative multiple imputation techniques for handling missing data were implemented using the sklearn. tree and sklearn.experimental packages. Functions for statistical testing and distribution analysis were provided by the scipy.stats package. Mediation analysis was performed using the pingouin package, and logistics regression was conducted using the statsmodels package. Statistical significance was considered at a threshold of P < 0.05.

## Results

### Population characteristics

A total of 375,024 participants were involved in this study, among whom 107991 had IUTE. As shown in Table 1, compared to participants without IUTE, those with exposure were more likely to have a slightly younger median age, higher BMI and higher proportions of males, white race, smokers, and frequent drinkers. They also tended to have higher levels of blood LDL-c and triglycerides, as well as lower levels of HDL-c.

**Table 1.**
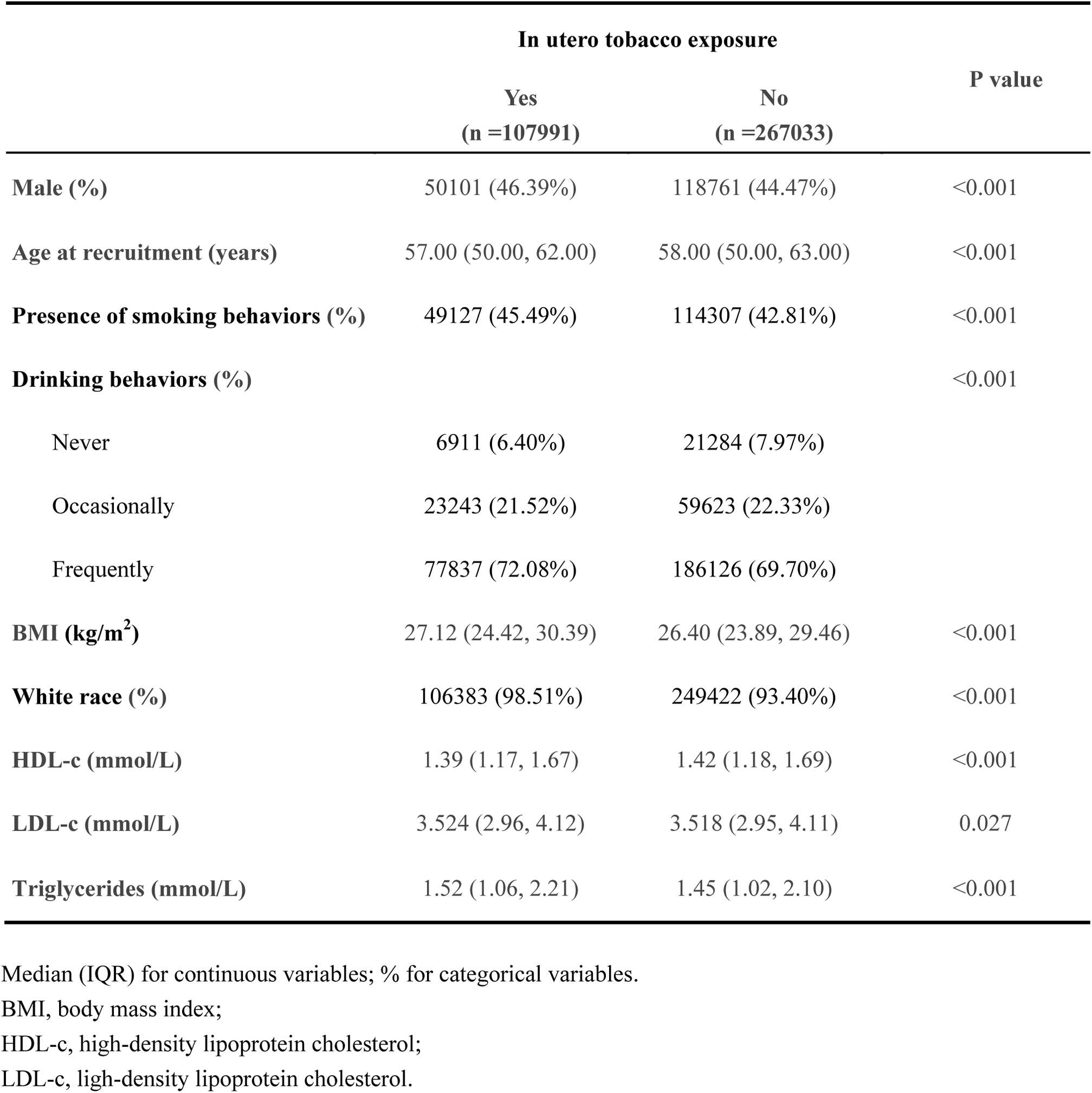
Baseline population characteristics of participants.

### IUTE and the risk of CVD

During a median follow-up period of 14.6 years, we identified 50,434 cases of CVD. Observed in the Kaplan-Meier survival curves, except for stroke (Log-rank p = 0.12) and nonrheumatic aortic valve disorders (Log-rank p = 0.57), the overall incidence rates of various CVDs and all-cause mortality over time were significantly higher in the IUTE group compared to the non-exposure group (Figure 2, Figure 3, Supplementary Figure 1-8). As shown in Table 2, after adjusting for all potential confounders, Cox regression analysis results revealed that IUTE significantly increased the risk of CVD incidence(HR 1.10, 95% CI 1.08-1.12), including myocardial infarction (HR 1.12, 95% CI 1.08-1.17), stroke (HR 1.07, 95% CI 1.02-1.13), atherosclerosis (HR 1.23, 95% CI 1.11-1.36), chronic ischemic heart disease (HR 1.08, 95% CI 1.05-1.11), heart failure (HR 1.09, 95% CI 1.04-1.14) and aortic aneurysm and dissection (HR 1.21, 95% CI 1.11-1.32). However, compared to the non-exposure group, the increased risks of nonrheumatic aortic valve disorders (P = 0.19) and cardiomyopathy (P = 0.12) were not significant in the IUTE group. Additionally, IUTE was significantly associated with a higher risk of all-cause mortality, which remained consistent across three models (Table 3; HR 1.07, 95% CI 1.05-1.10, model 1; HR 1.13, 95% CI 1.10-1.16, model 2; HR 1.11, 95% CI 1.09-1.14, model 3).

**Figure 2.**
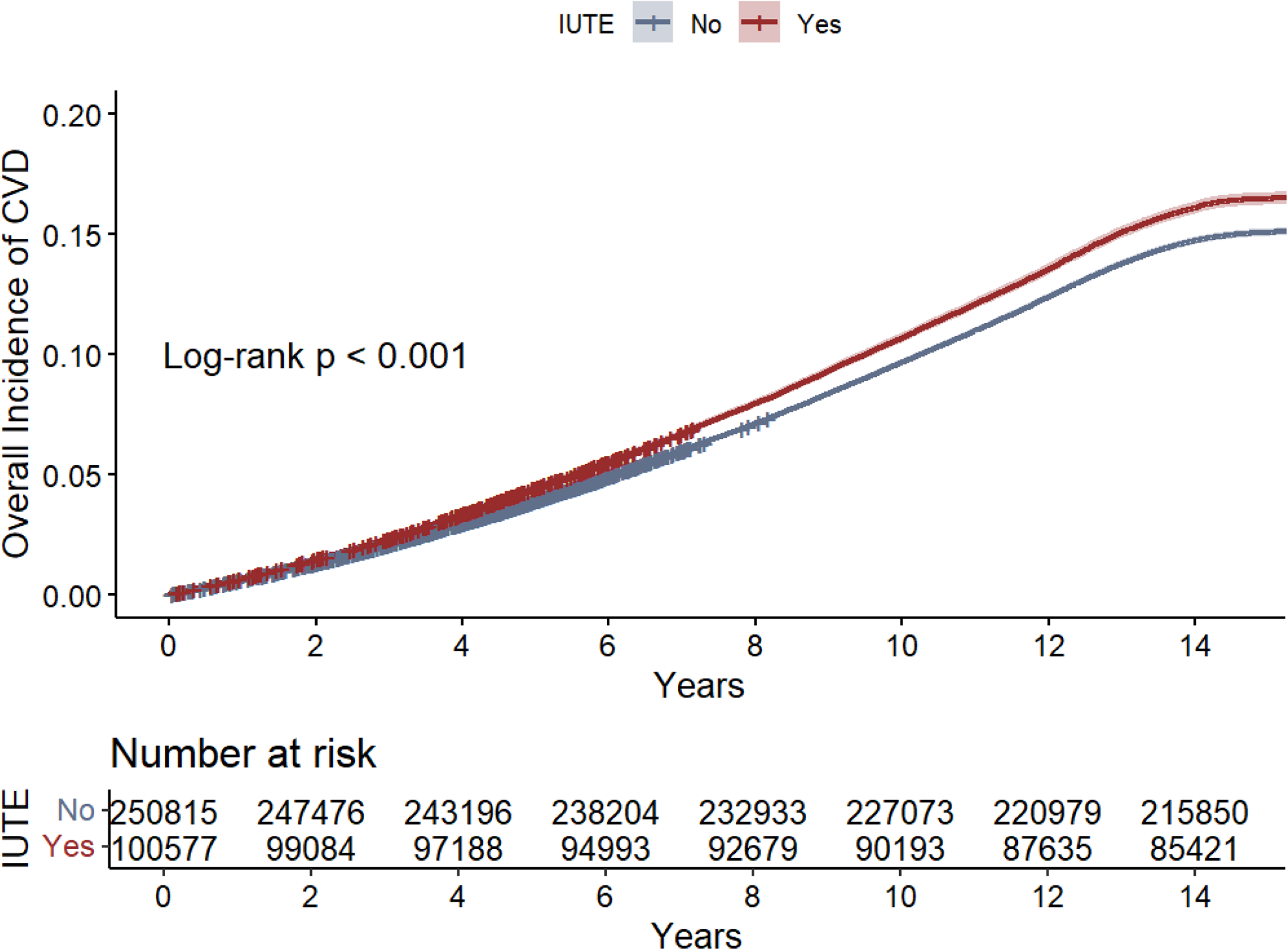
Overall incidence of CVD in groups with and without IUTE. The solid lines represent the estimated survival curves for different groups. The shaded area around these curves represent the 95% confidence intervals.

**Figure 3.**
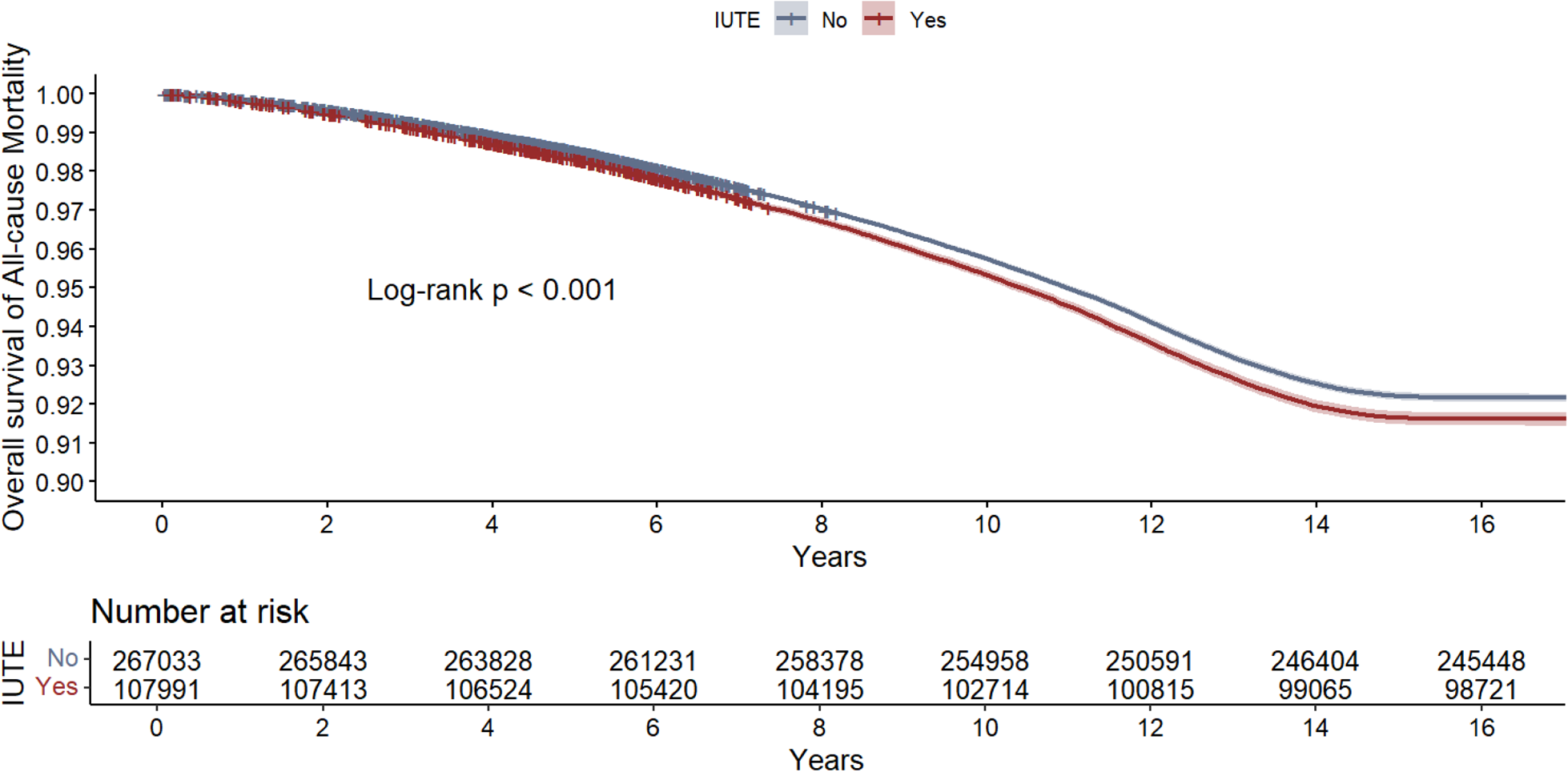
Overall incidence of all-cause mortality in groups with and without IUTE. The solid lines represent the estimated survival curves for different groups. The shaded area around these curves represent the 95% confidence intervals.

**Table 2.**
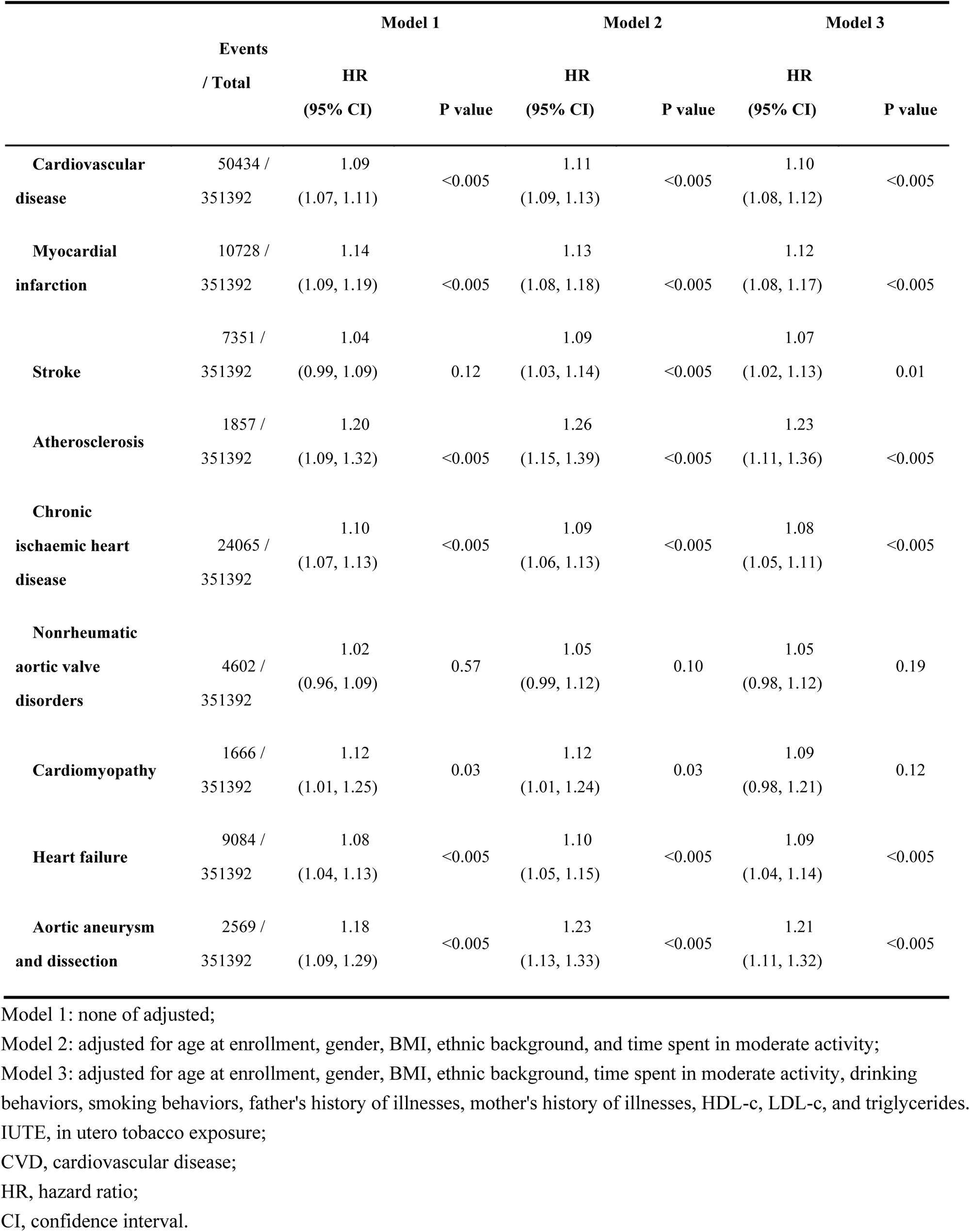
Association between IUTE and the risk of CVD.

**Table 3.** Association between IUTE and all-cause mortality.

|  | Model 1 | Model 2 | Model 3 |
| --- | --- | --- | --- |
| All-cause mortality |  |  |  |
| HR (95% CI) | 1.07 (1.05, 1.10) | 1.13 (1.10, 1.16) | 1.11 (1.09, 1.14) |
| P value | <0.005 | <0.005 | <0.005 |

Model 1: none of adjusted; Model 2: adjusted for age at enrollment, gender, BMI, ethnic background, and time spent in moderate activity; Model 3: adjusted for age at enrollment, gender, BMI, ethnic background, time spent in moderate activity, drinking behaviors, smoking behaviors, father's history of illnesses, mother's history of illnesses, HDL-c, LDL-c, and triglycerides. IUTE, in utero tobacco exposure; HR, hazard ratio; CI, confidence interval.

### IUTE, smoking behaviors after birth, and the risk of CVD

We observed a significant interaction effect (P for interaction < 0.005) between IUTE and smoking behaviors after birth on the risk of CVD. Regardless of smoking behaviors, IUTE significantly increased the incidence of CVD. Specifically, among individuals who smoked, IUTE was associated with a higher risk of myocardial infarction (HR 1.19, 95% CI 1.12-1.25), stroke (HR 1.12, 95% CI 1.04-1.20), atherosclerosis (HR 1.35, 95% CI 1.20-1.52), and heart failure (HR 1.15, 95% CI 1.08-1.22). The association between IUTE and chronic ischemic heart disease was significant in both individuals with and without smoking behaviors. It is worth noting that no interaction effect of before and after birth tobacco exposure was observed in outcome events such as nonrheumatic aortic valve disorders (P for interaction = 0.68), cardiomyopathy (P for interaction = 0.12), and aortic aneurysm and dissection (P for interaction = 0.42). These results are presented in Table 4.

**Table 4.** Association between IUTE and the risk of CVD stratified by smoking behaviors after birth.

|  | With smoking behaviors after birth |  | Without smoking behaviors after birth |  | P for interaction |
| --- | --- | --- | --- | --- | --- |
|  | HR (95% CI) | P value | HR (95% CI) | P value |  |
| **Cardiovascular disease** | 1.12 (1.09, 1.15) | <0.005 | 1.07 (1.04, 1.10) | <0.005 | <0.005 |
| **Myocardial infarction** | 1.19 (1.12, 1.25) | <0.005 | 1.04 (0.98, 1.11) | 0.22 | <0.005 |
| **Stroke** | 1.12 (1.04, 1.20) | <0.005 | 1.02 (0.94, 1.10) | 0.62 | 0.03 |
| **Atherosclerosis** | 1.35 (1.20, 1.52) | <0.005 | 0.99 (0.81, 1.19) | 0.88 | 0.01 |
| **Chronic ischaemic heart disease** | 1.10 (1.06, 1.14) | <0.005 | 1.05 (1.01, 1.09) | 0.03 | 0.04 |
| **Nonrheumatic aortic valve disorders** | 1.02 (0.93, 1.12) | 0.67 | 1.07 (0.98, 1.18) | 0.15 | 0.68 |
| **Cardiomyopathy** | 1.15 (0.99, 1.34) | 0.06 | 1.02 (0.87, 1.19) | 0.83 | 0.12 |
| **Heart failure** | 1.15 (1.08, 1.22) | <0.005 | 1.02 (0.95, 1.09) | 0.67 | <0.005 |
| **Aortic aneurysm and dissection** | 1.18 (1.06, 1.31) | <0.005 | 1.26 (1.09, 1.46) | <0.005 | 0.42 |

Adjusted for age at enrollment, gender, BMI, ethnic background, time spent in moderate activity, drinking behaviors, father's history of illnesses, mother's history of illnesses, HDL-c, LDL-c, and triglycerides.
IUTE, in utero tobacco exposure;
CVD, cardiovascular disease;
HR, hazard ratio;
CI, confidence interval.

### IUTE, PRS, and the risk of CVD

The study found a significant interaction between IUTE and PRS with CVD incidence (P for interaction < 0.005), indicating a positive association between IUTE and the risk of CVD across populations with different genetic susceptibilities (P < 0.005 for all). Specifically, the risk of stroke (HR 1.12, 95% CI 1.02-1.23), chronic ischemic heart disease (HR 1.10, 95% CI 1.05-1.16), cardiomyopathy (HR 1.31, 95% CI 1.07-1.60) and heart failure (HR 1.11, 95% CI 1.02-1.21) increased by IUTE was observed only in individuals with high genetic risk (fourth quartile). Among the four PRS quartiles, individuals with IUTE (compared to those without exposure) tended to have a higher risk of myocardial infarction. For atherosclerosis and aortic aneurysm and dissection, significant associations were observed only at higher (fourth quartile) and lower (first quartile) levels of genetic susceptibility. No significant association was found between IUTE and the risk of nonrheumatic aortic valve disorders across all levels of genetic susceptibility. Detailed results are presented in Table 5.

**Table 5.** Association between IUTE and the risk of CVD stratified by PRS for CVD.

|  | First quartile |  | Second quartile |  | Third quartile |  | Fourth quartile |  | P for interaction |
| --- | --- | --- | --- | --- | --- | --- | --- | --- | --- |
|  | HR (95% CI) | P value | HR (95% CI) | P value | HR (95% CI) | P value | HR (95% CI) | P value |  |
| **Cardiovascular disease** | 1.10 (1.05, 1.15) | <0.005 | 1.08 (1.04, 1.13) | <0.005 | 1.06 (1.02, 1.10) | <0.005 | 1.13 (1.09, 1.17) | <0.005 | <0.005 |
| **Myocardial infarction** | 1.16 (1.03, 1.30) | 0.01 | 1.14 (1.03, 1.25) | 0.01 | 1.09 (1.01, 1.19) | 0.03 | 1.11 (1.04, 1.19) | <0.005 | <0.005 |
| **Stroke** | 1.06 (0.94, 1.19) | 0.36 | 1.05 (0.94, 1.17) | 0.40 | 1.02 (0.92, 1.13) | 0.66 | 1.12 (1.02, 1.23) | 0.02 | <0.005 |
| **Atherosclerosis** | 1.32 (1.03, 1.69) | 0.03 | 1.11 (0.89, 1.37) | 0.36 | 1.16 (0.96, 1.41) | 0.12 | 1.34 (1.12, 1.61) | <0.005 | <0.005 |
| **Chronic ischaemic heart disease** | 1.05 (0.98, 1.13) | 0.19 | 1.06 (1.00, 1.13) | 0.06 | 1.05 (1.00, 1.11) | 0.06 | 1.10 (1.05, 1.16) | <0.005 | <0.005 |
| **Nonrheumatic aortic valve disorders** | 1.00 (0.86, 1.16) | 0.98 | 1.07 (0.93, 1.23) | 0.36 | 1.07 (0.94, 1.21) | 0.33 | 1.05 (0.92, 1.18) | 0.48 | <0.005 |
| **Cardiomyopathy** | 1.16 (0.93, 1.45) | 0.20 | 0.97 (0.77, 1.22) | 0.78 | 0.90 (0.72, 1.12) | 0.35 | 1.31 (1.07, 1.60) | 0.01 | 0.01 |
| **Heart failure** | 1.10 (0.99, 1.23) | 0.06 | 1.08 (0.98, 1.19) | 0.13 | 1.07 (0.98, 1.17) | 0.13 | 1.11 (1.02, 1.21) | 0.02 | <0.005 |
| **Aortic aneurysm and dissection** | 1.26 (1.04, 1.52) | 0.02 | 1.11 (0.93, 1.33) | 0.24 | 1.18 (1.00, 1.39) | 0.06 | 1.27 (1.08, 1.50) | <0.005 | <0.005 |

Adjusted for age at enrollment, gender, BMI, ethnic background, time spent in moderate activity, drinking behaviors, smoking behaviors, father's history of illnesses, mother's history of illnesses, HDL-c, LDL-c, and triglycerides.
IUTE, in utero tobacco exposure;
CVD, cardiovascular disease;
PRS, polygenic risk score;
HR, hazard ratio;
CI, confidence interval.

### IUTE and adverse behaviors

The cross-sectional study on IUTE and adverse behaviors after birth is discussed in Table 6. Results from fully adjusted models showed that compared to those without IUTE, individuals with such exposure had higher odds of smoking (OR 1.08, 95% CI 1.06-1.09) and drinking (OR 1.29, 95% CI 1.26-1.33).

**Table 6.** Associations between IUTE and adverse behaviors after birth (including smoking and drinking)

|  | Model 1 | Model 2 | Model 3 |
| --- | --- | --- | --- |
| **Smoking behaviors after birth** |  |  |  |
| OR (95% CI) | 1.12 (1.10, 1.13) | 1.09 (1.08, 1.11) | 1.08 (1.06, 1.09) |
| P value | <0.005 | <0.005 | <0.005 |
| **Drinking behaviors after birth** |  |  |  |
| OR (95% CI) | 1.27 (1.23, 1.30) | 1.30 (1.27, 1.34) | 1.29 (1.26, 1.33) |
| P value | <0.005 | <0.005 | <0.005 |

Model 1: none of adjusted;
Model 2: adjusted for BMI, ethnic background, and time spent in moderate activity;
Model 3: adjusted for BMI, ethnic background, time spent in moderate activity, drinking behaviors (only for the analysis of smoking), smoking behaviors (only for the analysis of drinking), HDL-c, and LDL-c.
IUTE, in utero tobacco exposure;
OR, odds ratio;
CI, confidence interval.

### Mediation effects of smoking behaviors and HDL-c on IUTE to CVD

The smoking behaviors after birth (Proportion = 12.40%, P < 0.001) and HDL-c(Proportion = 14.20%, P < 0.001) significantly mediated the relationship between IUTE and CVD risk (Figure 4). Specifically, when the outcome events were myocardial infarction, stroke, atherosclerosis, chronic ischaemic heart disease, heart failure, and aortic aneurysm and dissection, the corresponding mediation proportions (%) of smoking behaviors after birth in the associations were 11.70% (P < 0.001), 14.90% (P = 0.003), 14.30% (P < 0.001), 13.20% (P < 0.001), 14.50% (P < 0.001), and 16.30% (P = 0.001), respectively (Figure 5). As for HDL-c, the mediation proportions for the above diseases were 16.00% (P < 0.001), 12.70% (P = 0.001), 8.60% (P < 0.001), 18.50% (P < 0.001), 15.50% (P < 0.001), and 13.20% (P = 0.001), respectively (Figure 6). Since no significant association between IUTE and nonrheumatic aortic valve disorders (P = 0.19) or cardiomyopathy (P = 0.12) was found in the fully adjusted COX regression analysis (Table 2), here we haven’t performed mediation analysis for them.

**Figure 4.**
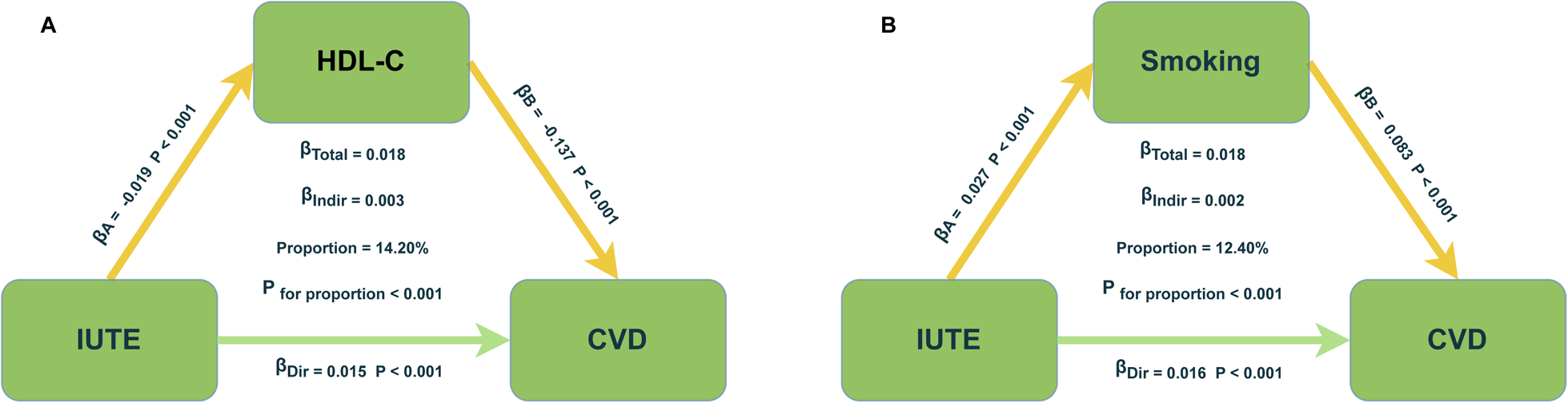
Mediation effects of smoking behaviors and HDL-c on IUTE to CVD. (A) The mediation role of HDL-c on IUTE to CVD; (B) The mediation role of smoking behaviors on IUTE to CVD.

**Figure 5.**
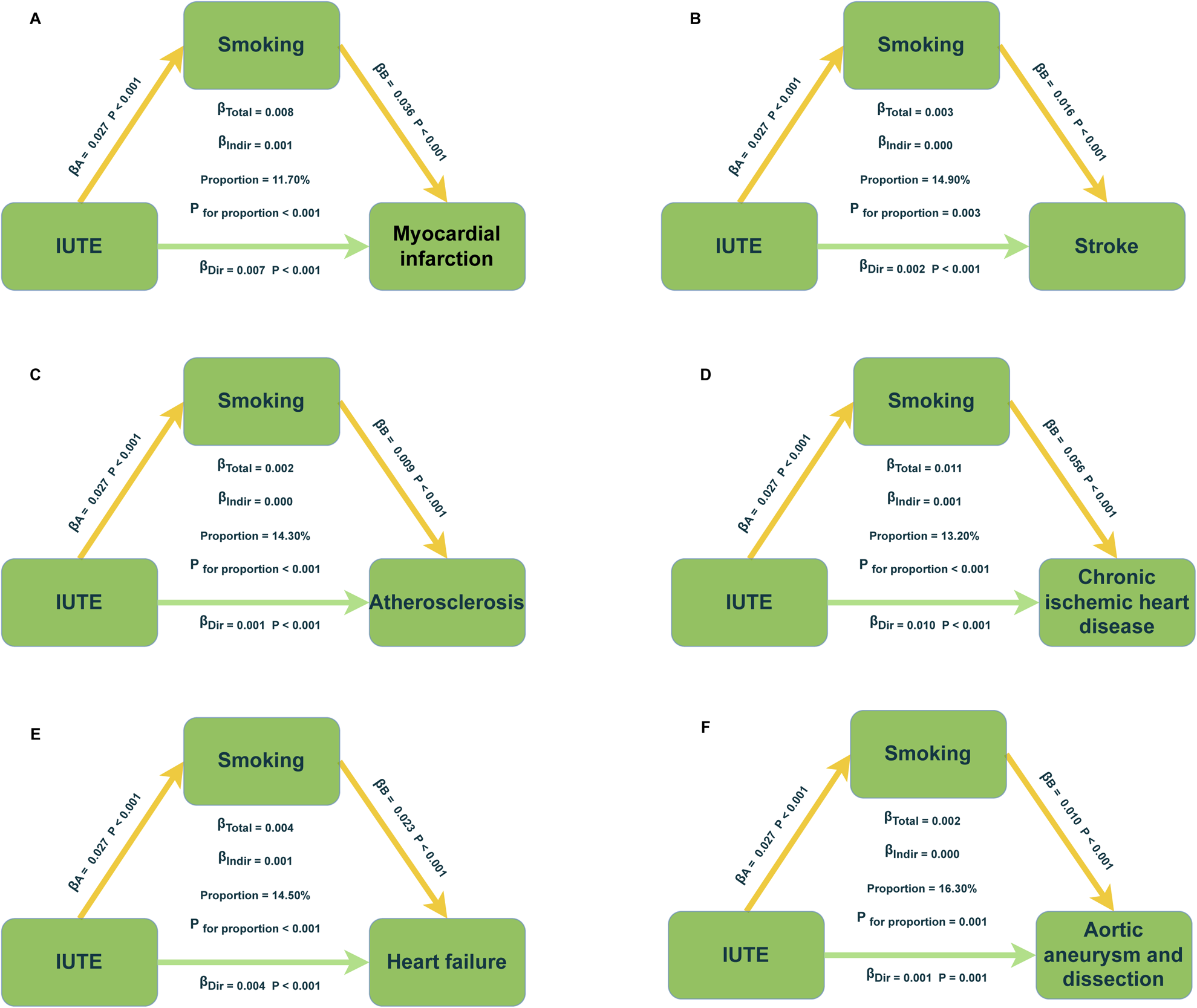
Mediation effects of smoking behaviors on IUTE to six clinically prevalent CVDs. (A) The mediation role of smoking behaviors on IUTE to myocardial infarction; (B) The mediation role of smoking behaviors on IUTE to stroke; (C) The mediation role of smoking behaviors on IUTE to atherosclerosis; (D) The mediation role of smoking behaviors on IUTE to chronic ischaemic heart disease; (E) The mediation role of smoking behaviors on IUTE to heart failure; (F) The mediation role of smoking behaviors on IUTE to aortic aneurysm and dissection.

**Figure 6.**
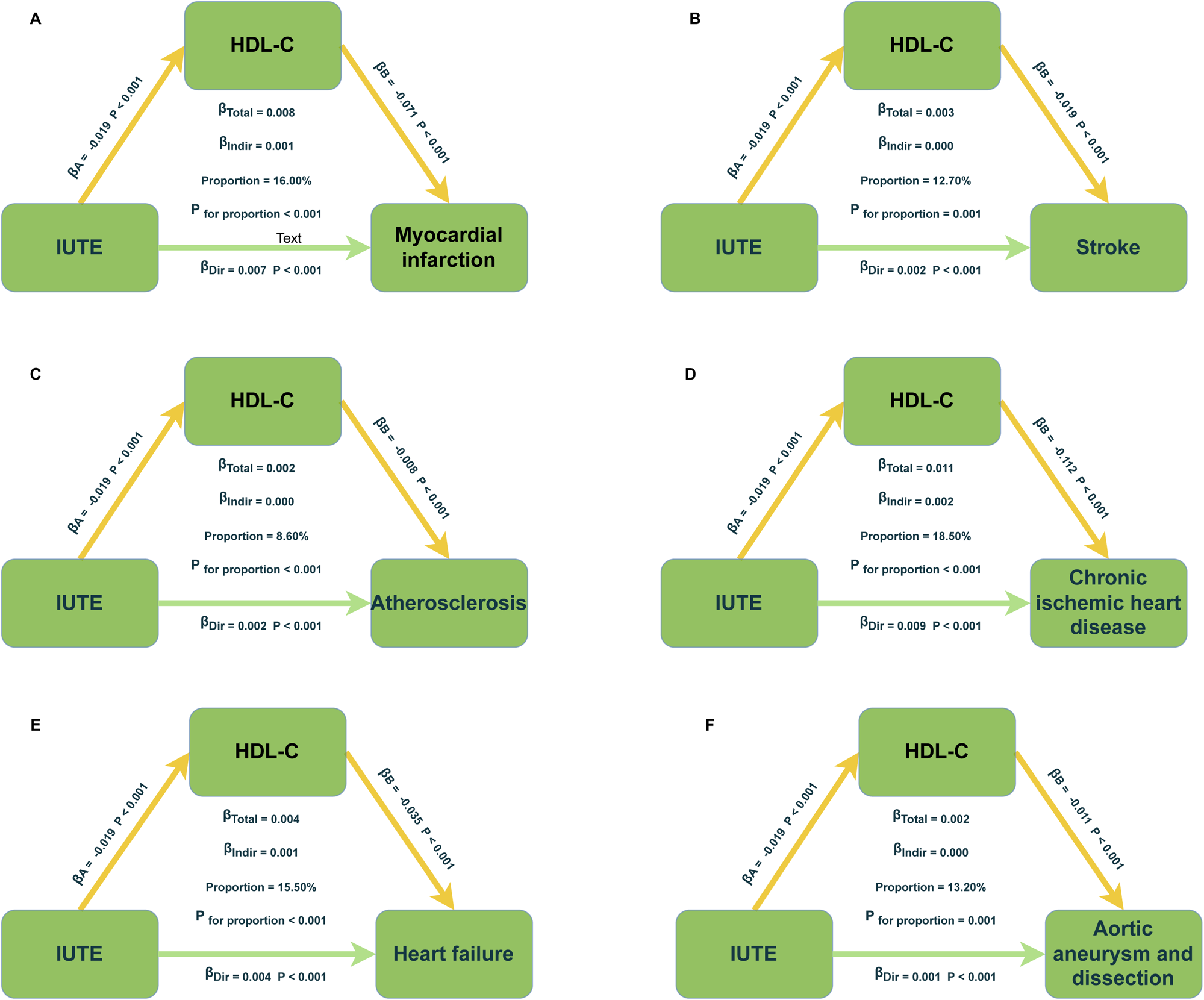
Mediation effects of HDL-c on IUTE to six clinically prevalent CVDs. (A) The mediation role of HDL-c on IUTE to myocardial infarction; (B) The mediation role of HDL-c on IUTE to stroke; (C) The mediation role of HDL-c on IUTE to atherosclerosis; (D) The mediation role of HDL-c on IUTE to chronic ischaemic heart disease; (E) The mediation role of HDL-c on IUTE to heart failure; (F) The mediation role of HDL-c on IUTE to aortic aneurysm and dissection.

## Discussion

In our population-based cohort study, which tracked participants over a 14.6-year follow-up period, we found evidence suggesting that IUTE might increase the risk of CVD in offspring. This association appears to be modulated by genetic predispositions and postnatal smoking behaviors. Intriguingly, our data also indicate that IUTE may heighten the propensity for smoking and alcohol consumption habits in offspring, potentially exacerbating their health risks. Moreover, we observed a significant rise in all-cause mortality rates among offspring exposed to IUTE. Through mediation analysis, we identified smoking behaviors and HDL-c levels as intermediaries in the link between IUTE and CVD, shedding light on possible mechanisms underlying these detrimental effects. Although previous studies have suggested a link between IUTE and increased CVD risk, our research provides the most comprehensive and thorough investigation of this connection to date.

The relationship between tobacco exposure and cardiovascular health has long been a subject of intense scrutiny. Our findings reveal that IUTE significantly raises the incidence of various cardiovascular diseases in adulthood, including myocardial infarction, stroke, atherosclerosis, chronic ischemic heart disease, heart failure, as well as aortic aneurysm and dissection. Consistent with our results, research by Song et al. has demonstrated that in utero exposure to maternal smoking is associated with an increased risk of myocardial infarction and stroke, even after adjusting for other traditional risk factors [28]. Additionally, our Cox regression results revealed that IUTE significantly increases all-cause mortality, an aspect previously unexplored in research. Previous studies have demonstrated that nicotine exposure exerts an influence on the vascular structures of the fetus [29–31]. The possible key factor is chemicals, including nicotine, free radicals, carbon monoxide, etc. may cause vasoconstriction, reduce placental blood flow, and impair placental and fetal vascular development [32]. Furthermore, scholarly investigations indicate that nicotine exposure during fetal development correlates with diminished concentrations of HDL in the offspring [33]. A study on rats exposed to nicotine (1 mg/kg/day) and lactation during pregnancy found abnormalities in the arrangement of the abdominal aortic wall and irregular arrangement of smooth muscle cells in the media. This irregular arrangement makes rats prone to forming atherosclerotic plaques [34]. In addition, a meta-analysis results indicate that smoking during pregnancy increases the risk of fibrillation [35], which is in line with our results. However, most of the contemporary studies concentrated on investigating the outcomes of maternal smoking or exposure to secondhand smoke during pregnancy on expectant mothers. For instance, Koba et al. have elucidated an elevated susceptibility to aortic aneurysm and dissection attributable to smoking [36]. To the best of our knowledge, comprehensive data and detailed in vivo or ex vivo studies that focus on the risk implications of tobacco exposure during pregnancy on offspring’s cardiovascular and cerebrovascular health remain scarce. Consequently, future research should prioritize exploring the underlying mechanisms of these disorders associated with IUTE.

We identified a significant interaction between IUTE and postnatal smoking behaviors in relation to the incidence of CVD. Consequently, we stratified the analysis based on smoking behavior post-birth. The results of this stratified analysis showed that individuals exposed to tobacco in utero and after birth had significantly increased risks of developing various CVDs, including myocardial infarction, stroke, atherosclerosis, chronic ischemic heart disease, cardiomyopathy, and heart failure. Previous literature has consistently emphasized the impact of parental smoking on the initiation of smoking behaviors during adolescence, thereby reinforcing the pivotal role of early tobacco exposure in shaping long-term smoking patterns and their attendant health implications [37]. Similarly, we found a significant interaction between IUTE and the Polygenic Risk Scores (PRS) for CVD in relation to the incidence of CVD. Stratified by quartiles of PRS, high genetic susceptibility, and IUTE together increased the risk of stroke, chronic ischemic heart diseases, cardiomyopathy, and heart failure. Additionally, even among individuals with lower genetic risk for CVD, significant associations were observed between IUTE and increased incidence of myocardial infarction, atherosclerosis, as well as aortic aneurysm and dissection. Previous research has discussed the combined impact of environmental factors and genetic risk on the occurrence of CVD [38–40], while our study provides evidence for the differential impact of IUTE on CVD risk across individuals with varying levels of genetic susceptibility. These findings highlight the necessity of adopting personalized approaches in CVD prevention and intervention, taking into account the dual influence of genetic and lifestyle factors.

Due to the scarcity of large-scale studies on the association between IUTE and the onset of smoking or other adverse behaviors, our cross-sectional study utilized logistic regression to examine the relationship between IUTE and the incidence of smoking and drinking behaviors. We found that IUTE increases the likelihood of smoking and drinking, consistent with previous findings. Many cohort studies have proved that IUTE increases the probability of smoking and alcohol abuse in offspring [41, 42], and shows a certain sex-specific prenatal effect, ie. maternal smoking during pregnancy had a positive effect on adolescent daughters’ drinking and smoking [43]. Therefore, we hypothesize that IUTE may increase the risk of CVD by promoting the onset of smoking and drinking behaviors.

A notable contribution of this study is the first-time discovery that an increase in the likelihood of smoking after birth and a decrease in HDL-c levels significantly mediate the relationship between IUTE and CVD risk. Previous research on IUTE has not delved deeply, merely investigating the association between IUTE and the incidence of smoking or the link with CVD risk, without exploring the potential mediating relationships. In the preceding sections, we clarified that IUTE increases the incidence of smoking behaviors, which are already well-established as significant risk factors for CVD [44]. Through mediation analysis, we further corroborated the hypothesis proposed earlier in our text. HDL-c measurement is one of the important predictors of CVD incidence, and many prospective studies from different racial and ethnic groups worldwide have confirmed that HDL-c is a strong, consistent, and independent predictor of the occurrence of cardiovascular events, including myocardial infarction, and ischemic stroke [45]. The mechanisms by which abnormalities of HDL-c contribute to various CVDs have been extensively studied. HDL-c is recognized as a heart-protective substance, which can promote the transport of cholesterol from peripheral tissues to the liver. It also has antioxidant and anti-inflammatory effects, such as reducing the risk of atherosclerosis by inhibiting LDL-c peroxidation, reducing the secretion of chemokines and cytokines to reduce inflammatory responses, etc [46]. Our findings suggest that IUTE reduces HDL-c levels, thereby diminishing its protective effect on the cardiovascular system, and consequently increasing the incidence of CVD among participants. Our study indicates that IUTE lowers HDL-c levels, diminishing its protective role against CVD and thereby increasing the incidence of CVD among our participants. Although the specific biological mechanisms driving these mediating effects are not yet fully understood, our findings offer new insights into the possible pathways through which IUTE heightens the risk of developing CVD. Considering the high prevalence of CVD in middle-aged and older populations, our results suggest that controlling smoking habits and managing HDL-c levels could be effective in reducing CVD risk, particularly among individuals exposed to tobacco in utero.

### Strengthens

Our study boasts several strengths. First, it encompasses a broad spectrum of CVDs and marks the first time the comprehensive relationship between CVD and IUTE has been investigated within a large cohort. Additionally, this is the inaugural study to assess the impact of IUTE on all-cause mortality. Second, aligning with previous findings that IUTE increases smoking behaviors, we executed a stratified analysis based on smoking behaviors, providing robust epidemiological evidence of IUTE’s detrimental effects on CVD risk. Unlike earlier studies, ours delved into genetic factors by stratifying participants based on the PRS for CVD, enabling a deeper analysis. Our comprehensive series of stratified analyses also substantiate the robustness of our findings. Furthermore, our research is pioneering in evaluating the mediating roles of smoking habits and HDL cholesterol levels in the increased risk of CVD linked to IUTE. It highlights how the onset of smoking habits and reductions in HDL-c levels may serve as critical pathways and biological mechanisms through which IUTE detrimentally impacts cardiovascular health.

### Limitations

There are several limitations to our study that warrant attention. Firstly, the UKB did not collect information on secondhand tobacco exposure for pregnant women, so the definition of IUTE in our study was limited to the mother’s smoking during pregnancy. However, it is undeniable that maternal smoking during pregnancy has the most direct impact on the fetus, thus our study remains robust. Future research should investigate whether secondhand tobacco exposure during pregnancy similarly affects offspring’s cardiovascular health. Secondly, data on maternal smoking were obtained from participants’ self-reports, decades after birth, which may introduce biases. Despite this, our study’s significant findings in a large cohort of over 300,000 participants provide valuable preliminary evidence. Thirdly, our results suggest that the correlation between IUTE and increased CVD risk is influenced by postnatal smoking behaviors. However, exposure to secondhand smoke during childhood, also likely among our cohort, may further elevate CVD risk, complicating our findings. Unfortunately, due to the lack of detailed data on such exposures in the UKB, these factors could not be accounted for. Fourthly, as reported by Fry et al., UKB participants typically reside in less socioeconomically deprived areas, which may indicate a ‘healthy volunteer’ bias[47]. Fifthly, since 96% of the UK Biobank volunteers are white British, our findings may not be generalizable to other ethnicities or the general population. Future studies should aim to replicate this research in more diverse populations. Despite these limitations, our meticulous data cleaning and robust analysis methods help ensure the reliability of our findings.

## Conclusion

In summary, our study indicates that IUTE increases the risk of CVD in adulthood, particularly among individuals who engage in smoking behaviors. It also raises the incidence of smoking and drinking behaviors after birth. Furthermore, we found that the impact of IUTE on CVD incidence is mediated by smoking behaviors after birth and HDL-c, and is influenced by genetic susceptibility to CVD. Additionally, IUTE was associated with increased all-cause mortality. These findings emphasize the potential adverse effects of maternal smoking habits during pregnancy on offspring, highlighting the necessity for public health education and regulation to address unhealthy behaviors like smoking during pregnancy. Further research should focus on the specific mechanisms through which IUTE influences CVD incidence.

## Author Contributions

Drs ZW Zhu, YX Zheng and JingBao had full access to all of the data in the study and take responsibility for the integrity of the data and the accuracy of the data analysis.

Concept and design: YX Zheng.

Acquisition, analysis, or interpretation of data: YX Zheng, J Bao, Y Guo.

Drafting of the manuscript: XY Xiong, ZX Chen, F Zou, J Bao, JY Liu, YX Zheng, J Wang, QY Wang, YX Qiu.

Critical revision of the manuscript for important intellectual content: ZW Zhu, YX Zheng, XY Xiong.

Statistical analysis: YX Zheng, JingBao.

Obtained funding: ZW Zhu.

Administrative, technical, or material support: XY Xiong, YX Zheng, J Bao.

Supervision: YX Zheng, ZW Zhu.

## Data Availability

The data are available from UKB Resource (www.ukbiobank.ac.uk/). This research has been conducted using the UKB Resource under application number 84709.

## Ethical Approval

The UKB study protocol was approved by the North West Multi-centre Research Ethics Committee.

## Pre-registered Clinical Trial Number

None supplied.

## Reference

1. Lloyd-Jones, D., et al., Executive summary: heart disease and stroke statistics--2010 update: a report from the American Heart Association. Circulation, 2010. 121(7): p. 948–54.

2. Global burden of 369 diseases and injuries in 204 countries and territories, 1990-2019: a systematic analysis for the Global Burden of Disease Study 2019. Lancet, 2020. 396(10258): p. 1204–1222.

3. Mukamal, K. and M. Lazo, Alcohol and cardiovascular disease. Bmj, 2017. 356: p. j1340.

4. Larsson, S.C., et al., Genetic predisposition to smoking in relation to 14 cardiovascular diseases. Eur Heart J, 2020. 41(35): p. 3304–3310.

5. Bianchi, C., et al., Primary prevention of cardiovascular disease in people with dysglycemia. Diabetes Care, 2008. 31 Suppl 2: p. S208–14.

6. Khoury, M. and E.M. Urbina, Hypertension in adolescents: diagnosis, treatment, and implications. Lancet Child Adolesc Health, 2021. 5(5): p. 357–366.

7. Barker, D.J., Fetal origins of coronary heart disease. Bmj, 1995. 311(6998): p. 171–4.

8. Perak, A.M., et al., Associations of Maternal Cardiovascular Health in Pregnancy With Offspring Cardiovascular Health in Early Adolescence. Jama, 2021. 325(7): p. 658–668.

9. Reynolds, R.M., et al., Maternal obesity during pregnancy and premature mortality from cardiovascular event in adult offspring: follow-up of 1 323 275 person years. Bmj, 2013. 347: p. f4539.

10. Yuan, X., et al., Maternal Exposure to PM(2.5) and the Risk of Congenital Heart Defects in 1.4 Million Births: A Nationwide Surveillance-Based Study. Circulation, 2023. 147(7): p. 565–574.

11. Lange, S., et al., National, regional, and global prevalence of smoking during pregnancy in the general population: a systematic review and meta-analysis. Lancet Glob Health, 2018. 6(7): p. e769–e776.

12. Toppila-Salmi, S., et al., Maternal smoking during pregnancy affects adult onset of asthma in offspring: a follow up from birth to age 46 years. Eur Respir J, 2020. 55(6).

13. Cui, F., et al., Early-life exposure to tobacco, genetic susceptibility, and accelerated biological aging in adulthood. Sci Adv, 2024. 10(18): p. eadl3747.

14. Peng, Y., et al., Maternal smoking during pregnancy increases the risk of gut microbiome-associated childhood overweight and obesity. Gut Microbes, 2024. 16(1): p. 2323234.

15. Zou, R., et al., Association of Maternal Tobacco Use During Pregnancy With Preadolescent Brain Morphology Among Offspring. JAMA Netw Open, 2022. 5(8): p. e2224701.

16. Corrêa, M.L., et al., Maternal smoking during pregnancy and children’s mental healthat age 22 years: Results of a birth cohort study. J Affect Disord, 2022. 300: p. 203–208.

17. Nogueira Avelar, E.S.R., et al., Associations of Maternal Diabetes During Pregnancy With Psychiatric Disorders in Offspring During the First 4 Decades of Life in a Population-Based Danish Birth Cohort. JAMA Netw Open, 2021. 4(10): p. e2128005.

18. Hackshaw, A., C. Rodeck, and S. Boniface, Maternal smoking in pregnancy and birth defects: a systematic review based on 173 687 malformed cases and 11.7 million controls. Hum Reprod Update, 2011. 17(5): p. 589–604.

19. de Simone, G., et al., Hypertension in children and adolescents. Eur Heart J, 2022. 43(35): p. 3290–3301.

20. Taylor, K., et al., The effect of maternal BMI, smoking and alcohol on congenital heart diseases: a Mendelian randomisation study. BMC Med, 2023. 21(1): p. 35.

21. Wang, T., et al., Maternal pre-pregnancy/early-pregnancy smoking and risk of congenital heart diseases in offspring: A prospective cohort study in Central China. J Glob Health, 2022. 12: p. 11009.

22. Bolin, E.H., et al., Maternal Smoking and Congenital Heart Defects, National Birth Defects Prevention Study, 1997-2011. J Pediatr, 2022. 240: p. 79–86.e1.

23. Taylor, K., et al., Effect of Maternal Prepregnancy/Early-Pregnancy Body Mass Index and Pregnancy Smoking and Alcohol on Congenital Heart Diseases: A Parental Negative Control Study. J Am Heart Assoc, 2021. 10(11): p. e020051.

24. Leybovitz-Haleluya, N., et al., Maternal smoking during pregnancy and the risk of pediatric cardiovascular diseases of the offspring: A population-based cohort study with up to 18-years of follow up. Reprod Toxicol, 2018. 78: p. 69–74.

25. Sudlow, C., et al., UK biobank: an open access resource for identifying the causes of a wide range of complex diseases of middle and old age. PLoS Med, 2015. 12(3): p. e1001779.

26. Thompson, D.J., et al., UK Biobank release and systematic evaluation of optimised polygenic risk scores for 53 diseases and quantitative traits. medRxiv, 2022: p. 2022.06.16.22276246.

27. Lee, J. and K.P. Fung, Confidence interval of the kappa coefficient by bootstrap resampling. Psychiatry Res, 1993. 49(1): p. 97–8.

28. Song, Q., et al., Perinatal exposure to maternal smoking and adulthood smoking behaviors in predicting cardiovascular diseases: A prospective cohort study. Atherosclerosis, 2021. 328: p. 52–59.

29. Bakker, H. and V.W. Jaddoe, Cardiovascular and metabolic influences of fetal smoke exposure. Eur J Epidemiol, 2011. 26(10): p. 763–70.

30. Benedict, L.M., et al., Identifying and Preventing Cardiac Risk Factors from Fetal Life. Journal of Pediatrics and Pediatric Medicine, 2018. 2(3).

31. Mishra, A., et al., Harmful effects of nicotine. Indian Journal of Medical and Paediatric Oncology, 2015. 36(01): p. 24–31.

32. Lambers, D.S. and K.E. Clark, The maternal and fetal physiologic effects of nicotine. Seminars in Perinatology, 1996. 20(2): p. 115–126.

33. Turan, H., et al., Assessment of carotid artery intima-media thickness and lipid profile in term neonates born to smoking mothers. Iranian Journal of Pediatrics, 2021.

34. Alfourti, A., A. Azzwali, and A. Azab, Effects of Maternal Nicotine Exposure during Pregnancy and Lactation on Blood Pressure of the Offspring and Blood Vessel Structure and Attenuation by Vitamin C. East Afr. Sch. J. Med. Sci, 2019. 2: p. 240–246.

35. Aune, D., et al., Tobacco smoking and the risk of atrial fibrillation: A systematic review and meta-analysis of prospective studies. Eur J Prev Cardiol, 2018. 25(13): p. 1437–1451.

36. Koba, A., et al., Risk Factors for Mortality From Aortic Aneurysm and Dissection: Results From a 26-Year Follow-Up of a Community-Based Population. J Am Heart Assoc, 2023. 12(8): p. e027045.

37. Alves, J., et al., Intergenerational transmission of parental smoking: when are offspring most vulnerable? Eur J Public Health, 2022. 32(5): p. 741–746.

38. Zhang, J., et al., Relation of Life’s Essential 8 to the genetic predisposition for cardiovascular outcomes and all-cause mortality: results from a national prospective cohort. Eur J Prev Cardiol, 2023. 30(15): p. 1676–1685.

39. Diao, T., et al., Changes in Sleep Patterns, Genetic Susceptibility, and Incident Cardiovascular Disease in China. JAMA Netw Open, 2024. 7(4): p. e247974.

40. Zhang, H., et al., Familial factors, diet, and risk of cardiovascular disease: a cohort analysis of the UK Biobank. Am J Clin Nutr, 2021. 114(5): p. 1837–1846.

41. Al Mamun, A., et al., Does maternal smoking during pregnancy predict the smoking patterns of young adult offspring? A birth cohort study. Tob Control, 2006. 15(6): p. 452–7.

42. Nomura, Y., S.E. Gilman, and S.L. Buka, Maternal Smoking During Pregnancy and Risk of Alcohol Use Disorders Among Adult Offspring. Journal of Studies on Alcohol and Drugs, 2011. 72(2): p. 199–209.

43. Ncube, C.N. and B.A. Mueller, Daughters of Mothers Who Smoke: A Population-based Cohort Study of Maternal Prenatal Tobacco use and Subsequent Prenatal Smoking in Offspring. Paediatr Perinat Epidemiol, 2017. 31(1): p. 14–20.

44. Münzel, T., et al., Effects of tobacco cigarettes, e-cigarettes, and waterpipe smoking on endothelial function and clinical outcomes. Eur Heart J, 2020. 41(41): p. 4057–4070.

45. Toth, P.P., et al., High-density lipoproteins: a consensus statement from the National Lipid Association. J Clin Lipidol, 2013. 7(5): p. 484–525.

46. Säemann, M.D., et al., The versatility of HDL: a crucial anti-inflammatory regulator. Eur J Clin Invest, 2010. 40(12): p. 1131–43.

47. Fry, A., et al., Comparison of Sociodemographic and Health-Related Characteristics of UK Biobank Participants With Those of the General Population. Am J Epidemiol, 2017. 186(9): p. 1026–1034.

